# Theta deep-brain stimulation improves cognitive performance in Parkinson’s patients with cognitive impairments

**DOI:** 10.64898/2026.03.12.26348246

**Authors:** Rachel C. Cole, James F. Cavanagh, Qiang Zhang, Christopher L. Groth, Juan Vivanco-Suarez, Arturo I. Espinoza, Jeremy D. Greenlee, Nandakumar S Narayanan

**Affiliations:** Department of Neurology, University of Minnesota; Department of Psychology, University of New Mexico, Albuquerque, New Mexico, 87131; Department of Neurology, University of Iowa, Iowa City, IA, 52242; Des Moines University, West Des Moines, IA 50266; Department of Neurosurgery, University of Iowa, Iowa City, IA, 52242

## Abstract

**Background:** Patients with Parkinson’s disease (PD) almost inevitably experience cognitive impairments. These deficits have been linked to low frequency “theta” cortical activity ∼4 Hz, previously associated with cognitive control.

**Objective:** Our study investigated effects of 4 Hz subthalamic nucleus (STN) deep brain stimulation (DBS) on cognitive performance in PD patients with cognitive impairments.

**Methods:** We recruited 17 PD participants with (n=10) and without (n=7) cognitive impairment. In these patients, we compared motor and cognitive performance during 4 Hz STN DBS, typical DBS for motor symptoms of PD (∼130Hz) and DBS OFF. Motor performance was tested by Part III of the Movement Disorders Society Unified Parkinson’s Disease Rating Scale (MDS-UPDRS-III). Cognitive performance was tested during performance of the Multi-Source Interference Task (MSIT), which requires conflict resolution to respond accurately.

**Results:** Motor function improved with 4 Hz STN DBS and further improved with ∼130 Hz STN DBS. Compared to DBS OFF, reaction times were decreased during 4 Hz STN DBS and were further decreased at ∼130 Hz. Strikingly, 4 Hz DBS alone improved accuracy compared to both DBS OFF and compared to ∼130 Hz STN DBS.

**Conclusions:** These data suggest that theta-frequency 4 Hz STN stimulation is effective in PD patients with cognitive impairments. Our findings will help guide new therapies targeted at improving cognitive dysfunction in PD and could broaden applications for low-frequency brain stimulation.

## INTRODUCTION

Parkinson’s disease (PD) is a movement disorder with devastating cognitive symptoms that primarily impair executive functions, such as working memory, reasoning, and cognitive control (Narayanan and Albin, 2022). PD-related cognitive impairment (PDCI) can emerge early in disease and can progress to mild-cognitive impairment (MCI) or dementia (PDD) (Aarsland et al., 2011, 2008; Aarsland and Kurz, 2010). Although cognitive dysfunction eventually affects >80% of patients with PD (Hely et al., 2008; Williams-Gray et al., 2006), there are few reliable treatments for these symptoms (Zhang et al., 2020). Patients report that cognitive symptoms are a major factor influencing their quality of life (Korczyn, 2001), and cognitive impairment may disqualify them from highly effective therapies for motor symptoms such as deep-brain stimulation (DBS) (Montgomery, 2014). There is an urgent need to establish new diagnostic tools and treatments for cognitive dysfunction in PDCI.

PDCI is linked to widespread cortical dysfunction and altered low-frequency cortical activity in theta bands ∼4 Hz (Anjum et al., 2024; Caviness et al., 2015; Singh et al., 2023). Cortical theta activity is an established mechanism for cognitive control (Cavanagh and Cohen, 2022; Cavanagh and Frank, 2014; Yeager et al., 2024), suggesting that this system is particularly affected by PCDI (Yeager et al., 2024). Patients with PD have reduced evoked ∼4 Hz rhythms during interval timing, conflict, working memory, and reaction-time tasks (Cole et al., 2021; Singh et al., 2018, 2013; Solís-Vivanco et al., 2018, 2015; Ye et al., 2022; Yıldırım et al., 2024), and these deficits are predictive of PDCI (Anjum et al., 2024; Narayanan et al., 2024; Singh et al., 2023). Together, these findings provide robust motivation to target frontal theta activity in order to boost executive function in PD.

Cortical structures can be modulated through subthalamic nucleus (STN) deep brain stimulation (DBS) through antidromic stimulation of monosynaptic hyperdirect connections and/or feedforward projections through the thalamus (Chen et al., 2020; Kelley et al., 2018; Nambu et al., 2002; Walker et al., 2012; Xie et al., 2025). Intraoperative recordings have shown that theta oscillations synchronize frontal areas with the STN (Kelley et al., 2018; Zavala et al., 2016, 2014). Typical STN DBS is set to high frequencies ∼130 Hz to treat motor symptoms of PD, but can be set to much lower frequencies, for example ∼4 Hz (Okun, 2012). STN DBS at these lower frequencies does not target motor symptom improvement, but does show improvements in a variety of control tasks, including interval timing, Stroop performance, working memory, and reaction-time decision thresholds (Cole et al., 2025; Kelley et al., 2018; Salehi et al., 2024; Scangos et al., 2018; Xie et al., 2025). Together, these findings motivate the hypothesis that increasing low-frequency STN rhythms improves cognitive performance in patients with PD. In this report, we examined whether this intervention was specifically effective in patients with PDCI.

We tested this hypothesis in 17 PD patients with and without cognitive impairments implanted with STN DBS. We used the Multi-Source Interference Task (MSIT) in which responses require resolution of instructional interference, distractors, and spatial conflict (Bush et al., 2003; Herman et al., 2023; Sklar et al., 2017). We compared MSIT performance at 4 Hz STN DBS with stimulation frequencies optimized for PD motor signs (∼130 Hz) and DBS OFF conditions. Our findings provide evidence that 4 Hz DBS improves MSIT performance specifically in patients with PDCI, and therefore may inform future treatment strategies and diagnostic tools targeted at improving cognitive deficits in PD.

## MATERIALS AND METHODS

### Participants

PD participants were recruited from the movement disorders clinics at the University of Iowa. Twenty individuals with PD and bilateral STN DBS participated in this study (Table 1). Participants were required to have been treated with bilateral STN DBS for at least 6 months and to be on stable DBS settings. These are typically parameters effective for best motor function. All experimental procedures were approved by the University of Iowa Institutional Review Board (IRB # 201707828). All participants provided written informed consent after thoroughly reviewing the consent document with the experimenter. Participant demographics, medications, and DBS settings are shown in Table 1.

**Table 1:**
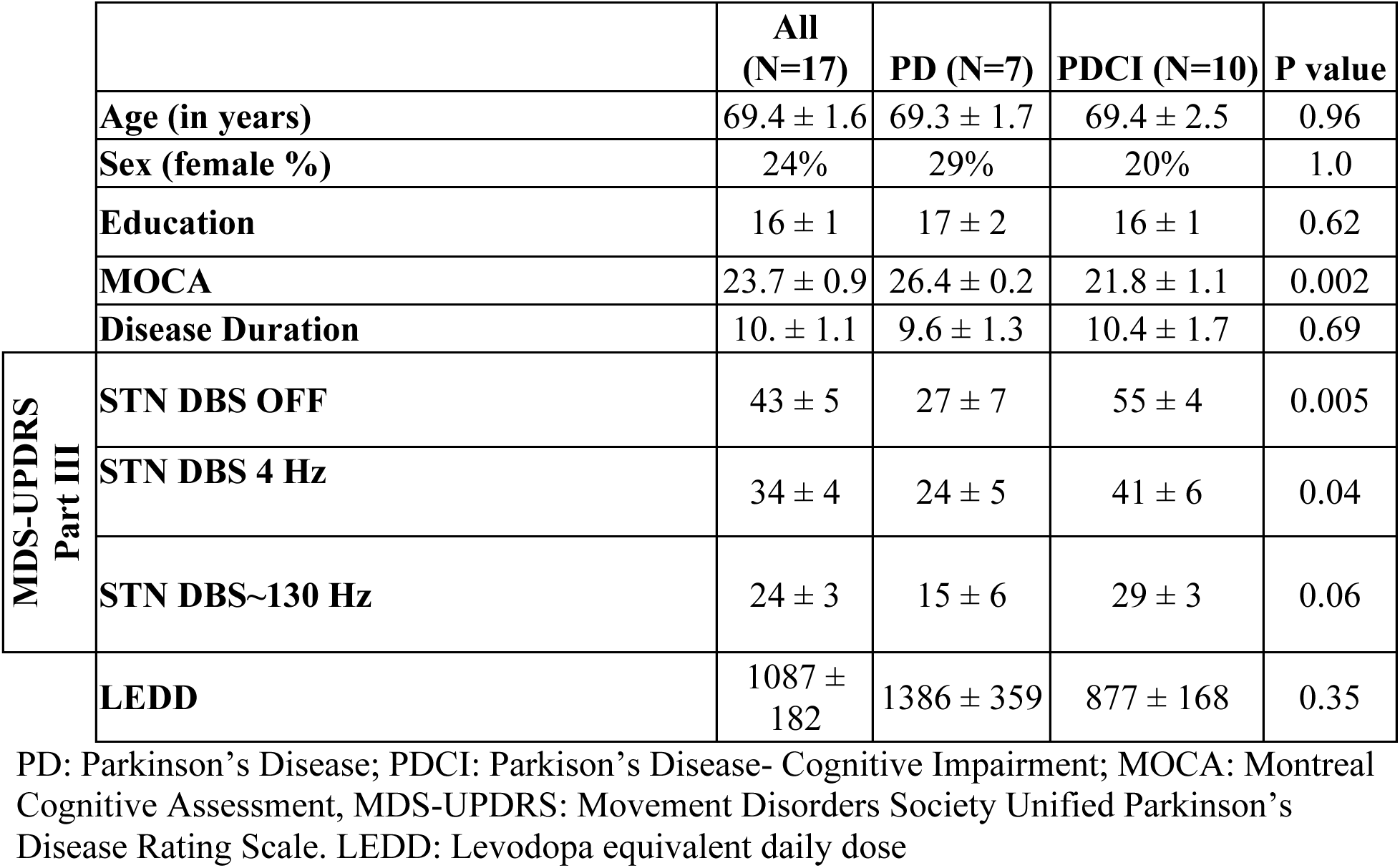
Demographic summary for 27 PD STN DBS patients who completed the MSIT.

|  |  | All<br>(N=17) | PD (N=7) | PDCI (N=10) | P value |
| --- | --- | --- | --- | --- | --- |
|  | Age (in years) | 69.4 ± 1.6 | 69.3 ± 1.7 | 69.4 ± 2.5 | 0.96 |
|  | Sex (female %) | 24% | 29% | 20% | 1.0 |
|  | Education | 16 ± 1 | 17 ± 2 | 16 ± 1 | 0.62 |
|  | MOCA | 23.7 ± 0.9 | 26.4 ± 0.2 | 21.8 ± 1.1 | 0.002 |
|  | Disease Duration | 10. ± 1.1 | 9.6 ± 1.3 | 10.4 ± 1.7 | 0.69 |
| MDS-UPDRS<br>Part III | STN DBS OFF | 43 ± 5 | 27 ± 7 | 55 ± 4 | 0.005 |
|  | STN DBS 4 Hz | 34 ± 4 | 24 ± 5 | 41 ± 6 | 0.04 |
|  | STN DBS~130 Hz | 24 ± 3 | 15 ± 6 | 29 ± 3 | 0.06 |
|  | LEDD | 1087 ± 182 | 1386 ± 359 | 877 ± 168 | 0.35 |

PD: Parkinson's Disease; PDCI: Parkinson's Disease- Cognitive Impairment; MOCA: Montreal Cognitive Assessment, MDS-UPDRS: Movement Disorders Society Unified Parkinson's Disease Rating Scale. LEDD: Levodopa equivalent daily dose

Participants were instructed to take medications as usual and to bring them to take during the session, if necessary. They were also asked to avoid caffeine and alcohol two hours prior to and during the session. Participants completed all tasks in a single testing session. During the session, participants completed a series of motor and cognitive tasks under three conditions: 1) while receiving bilateral STN DBS at 4 Hz; 2) while receiving bilateral STN DBS at their usual settings (80 – 160 Hz or ∼130 Hz; Table S1); 3) DBS OFF. During 4 Hz DBS, all other DBS parameters, including active electrode contacts, pulse width, and voltage were kept constant. Twenty participants were enrolled in the study; only 17 were able to complete the MSIT while receiving STN DBS under all three stimulation conditions and were included in the analysis. Some participants also participated in other cognitive tasks as detailed in (Cole et al., 2025).

We defined cognitive impairments in participants via the Montreal Cognitive Assessment (MOCA) administered on the day of testing (Dalrymple-Alford et al., 2010; Nasreddine et al., 2005; Singh et al., 2023). The MOCA score was used to separate PD patients into 2 groups based on cognitive status: patients who scored ≥26 were included in the group of PD without cognitive impairment (PD), and those who scored <26 were included in the group with cognitive impairment (PDCI; Dalrymple-Alford et al., 2010; Litvan et al., 2012). Within the PDCI group, MOCA scores ranged such that six participants were considered to have mild CI (PDMCI; MOCA 22-25) and four participants were considered to have PD dementia (PDD; MOCA<22) (Dalrymple-Alford et al., 2010; Litvan et al., 2012).

### Experimental procedures

All sessions began in the morning (typically ∼9 AM); each stimulation condition lasted about 75 minutes, and the entire experimental session lasted ∼4 hours. All experimental sessions were performed in a single day as in our prior work (Cole et al., 2025; Kelley et al., 2018; Singh et al., 2023). The experimental apparatus and procedures are detailed in Cole et al., 2025. Briefly, we used a Latin squares design to counterbalance the order of stimulation conditions to reduce order effects, which can be heavily influenced by fatigue, especially given the length of the session. The order of the stimulation conditions was either ∼130 Hz then 4 Hz then OFF, 4 Hz then OFF then ∼130 Hz, or OFF then ∼130 Hz then 4Hz. A movement disorders neurologist (N.N., C.G., or Q.Z.) performed all DBS adjustments and evaluated patients for adverse reactions. Stimulation changes were followed by a 20 minute washout period (Cooper et al., 2013). At the end of the washout period, we measured motor function by administering Part III of the MDS-UPDRS.

### Multi-Source Interference Task (MSIT)

The MSIT combines multiple dimensions of decision-making and was chosen because it involves high levels of task conflict (Fig 1). Participants had to press the corresponding keyboard keys with the index, middle, and ring fingers of their right hand: B if 1 was the target, N if 2 was the target, and M if 3 was the target. During *congruent* trials, distractor numbers were 0 and the target number was always in the position that was spatially congruent with the button press (e.g., if the number “1” appeared in the leftmost position (‘100’), participants had to press the letter ‘B’ with their index finger). By contrast, during *incongruent* trials the matching distractors were nonzero numbers and the unique target number was in a spatially incongruent position to the keyboard finger position (‘221’,’233’, etc.). Incongruent trials therefore required cognitive control to attend to the imperative cue and ignore distractors. Intertrial intervals randomly ranged between 500-1500ms. Participants were instructed to press the key that corresponded to the identity of the unique number, and in doing so, to ignore information about which position the numbers appeared in. Participants were instructed to respond as quickly and accurately as possible. After each trial, participants were given feedback on whether they were correct or incorrect and, if they were correct, whether they were faster or slower than the prior trial. If they were faster, the ‘correct’ text was colored in green. If the reaction time was slower, the ‘correct’ feedback text was red. Participants completed two blocks of 48 trials during each DBS stimulation condition over a total of 6 minutes. Within each block, each stimulus appeared an equivalent number of times. Participants were given a short, untimed break between blocks.

**Figure 1:**
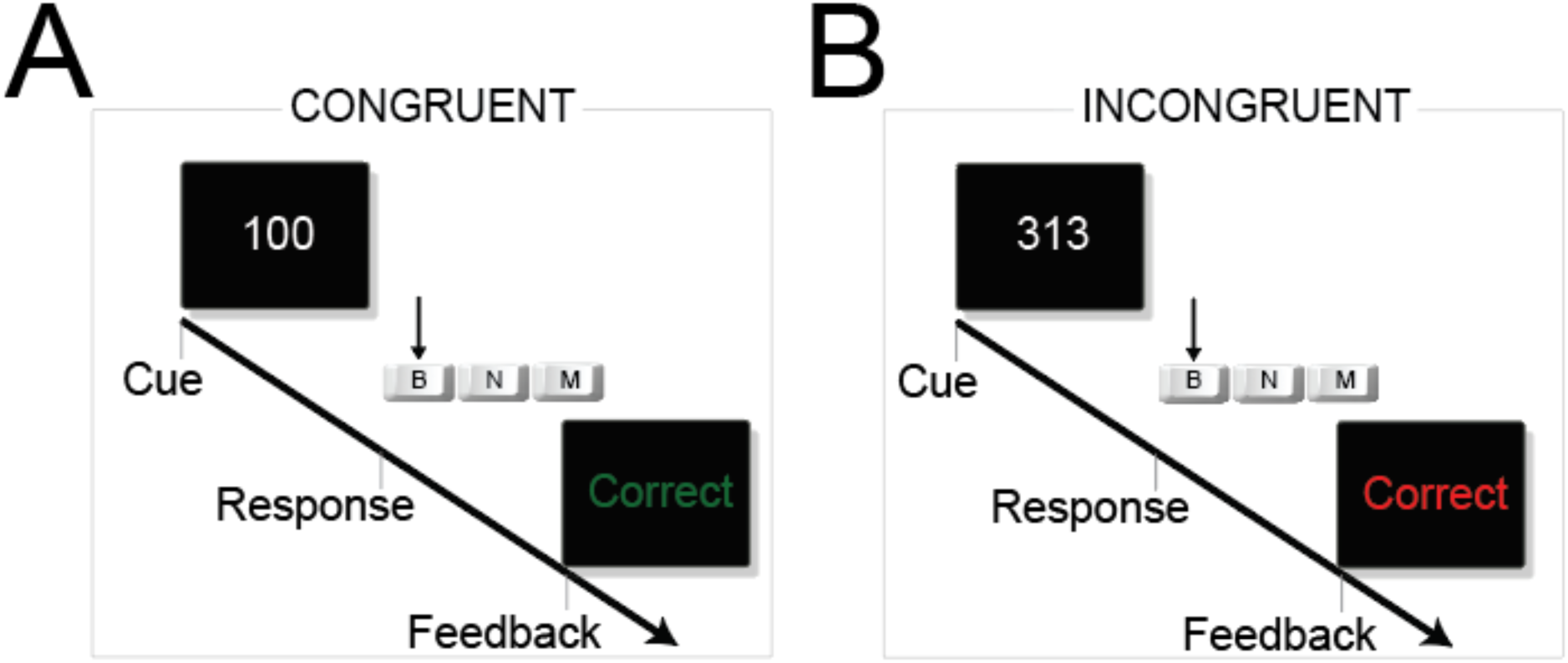
Multi-Source Interference Task (MSIT). **A)** Schematic of the MSIT for congruent and incongruent trials. During congruent trials, participants respond by pushing one of three keys on the left (B), middle (N), or right (M), as instructed by a 1, 2, or 3 respectively (e.g., participant would push “B” for the example shown). Other numbers are 0s. **B**) During incongruent trials, participants must determine the unique instructional cue (e.g., the 1 in the 313 sequence) and respond as instructed using the left, middle, or right key (e.g., B for the sequence shown). Incongruent trials have a high level of conflict in determining the instructions, ignoring distractors, and adjudicating spatial conflict. On all trials, participants received feedback: if the trial was correct, green text indicated a faster response time than the previous trial whereas red text indicated a slower response time. Incorrect feedback was presented as ‘incorrect’ in white.

Participants completed a short practice at the beginning of the research session consisting of detailed instructions, examples, and then a short block with 12 congruent trials followed by 12 incongruent trials. Participants received feedback on the screen after each trial, and the experimenter assisted with verbal cueing if the subject became confused or made many mistakes in a row. Key outcomes from the MSIT task were reaction times and accuracy.

### Statistical Analysis

All analyses were performed on trial-by-trial data via linear mixed effects models. Post-hoc testing was conducted using estimation marginal means (emmeans) with a false-discovery rate correction (Benjamini and Hochberg, 1995). A three-way interaction between cognitive status (PD vs. PDCI), stimulation condition (4 Hz, ∼130 Hz, and OFF), and trial type (Congruent vs. Incongruent) was considered for reaction time and accuracy models. For reaction times, we used a linear mixed effects model (*lmer* in R) on log-transformed data, and significance was evaluated using a Type III ANOVA with Satterthwaite-approximated degrees of freedom (*lmerTest* in R). To model the binomial distribution of trial-level accuracy, we used a generalized mixed-effects logistic regression model (*glmer* in R) and evaluated significance using Type III chi-square tests computed with the *car* package (*car::Anova*, type III in R).

We used the Akaike information criteria (AIC) values to select models, where a significant reduction of AIC values favors the model with more predictor variables. We found that including motor covariates MDS-UPDRS and LEDD did not improve our models significantly (reaction-time: ΔAIC = −3.9, χ²_(2)_ = 0.08, *p* = 0.96; accuracy: ΔAIC = −1.9, χ²_(2)_ = 5.9, *p* = 0.052), so these were omitted from the results presented here. Participant was included as a random effect in all models. All statistics were performed in close consultation with Biostatistics and Epidemiology Research and Design Core from the University of Iowa. Code and data are available at https://doi.org/10.5281/zenodo.18985728.

## RESULTS

We analyzed data from 17 patients who received STN DBS (Fig 2A) (Horn et al., 2019). Based on MOCA scores (Dalrymple-Alford et al., 2010; Litvan et al., 2012; Singh et al., 2023), seven were characterized as normal cognition (PD) and 10 with PDCI. Data for each participant including levodopa-equivalent dose and programming settings are shown in Table S1.

**Figure 2:**
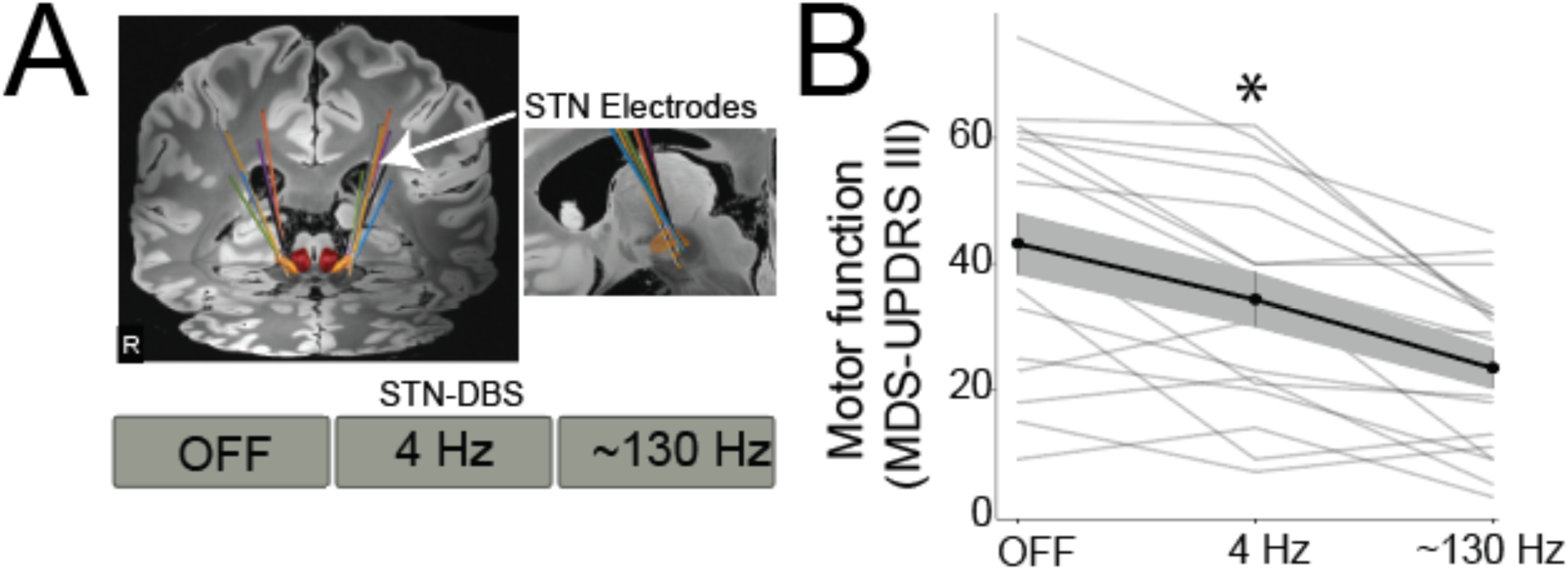
Subthalamic Nucleus (STN) Deep Brain Stimulation (DBS). **A**) Localization of STN electrodes in patients receiving DBS; LEAD-DBS localization data was available in 7 patients. **B**) Movement Disorders Society-Unified Parkinson’s Disease Rating Scale III (MDS-UPDRS-III) with STN DBS OFF, at 4 Hz, and at ∼130 Hz. *=main effect of STN DBS, suggesting that motor symptoms of PD improved at 4 Hz STN DBS and further improved with ∼130 Hz STN DBS. Data from 17 PD patients with STN DBS; thin lines represent mean data from a single patient; thick lines represent mean across patients, and shaded area represents standard error of the mean.

As with our past work (Cole et al., 2025), we found a highly reliable beneficial effect of high-frequency STN DBS on MDS-UPDRS III (Mean±SEM: OFF: 43 ± 4; 4 Hz: 34 ± 4; ∼130 Hz; 23 ± 3; F=26.1, p=0.0000001; Fig 2B). Post-hoc testing revealed lower MDS-UPDRS III scores for both active stimulation conditions, with DBS OFF conditions having higher MDS-UPDRS III scores than 4 Hz STN DBS (*p*=0.003) and ∼130 Hz STN (*p*<0.0001), and ∼4 Hz STN DBS with higher MDS-UPDRS III scores than ∼130 Hz (*p*=0.0006). Together, these data suggest that PD motor symptoms improved with 4 Hz STN DBS and further improved at ∼130 Hz STN DBS.

We investigated cognitive performance using the MSIT (Fig 1C-D). We harnessed mixed-effects logistic regression to analyze how log-transformed trial-level reaction-time data was affected by cognitive status (PD vs PDCI), stimulation condition (STN DBS OFF, 4, or ∼130 Hz), and trial type (congruent vs. incongruent MSIT trials; Fig 2). As expected, we observed a main effect of trial type (*F*_(1, 4024)_ = 842.6, *p* < 0.0001) and a main effect of stimulation condition (*F*_(2, 4023)_ = 17.4, *p* < 0.0001), but not cognitive status, indicating that patients with PDCI had similar overall reaction times as patients without cognitive impairment. Crucially, there was a significant interaction between cognitive status and trial type (*F*_(1, 4024)_ = 4.2, *p* = 0.04), and between cognitive status and stimulation condition (*F*_(2, 4023)_ = 25.6, *p* < 0.0001). There was no interaction between stimulation condition and trial type, likely due to variability between participants (*F*_(2, 4023)_ = 2.2, *p* =0.11; Fig 3B). Finally, there was no three-way interaction (*F*_(2, 4023)_ = 0.1, *p* =0.87, Fig 3A).

**Figure 3:**
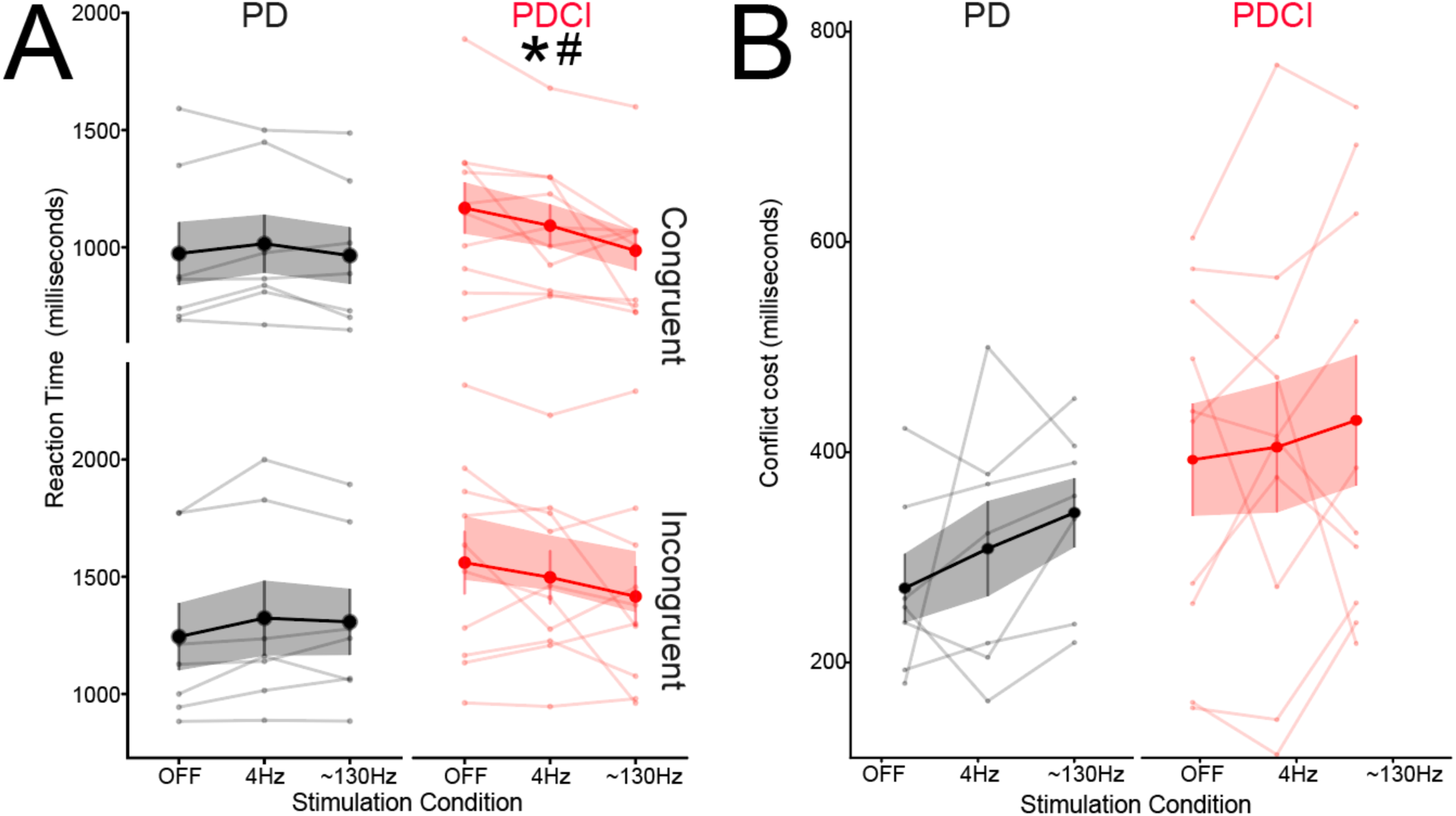
Subthalamic Nucleus (STN) Deep Brain Stimulation (DBS) at 4 Hz speeds reaction times in patients with PD Cognitive Impairment (PDCI). **A**) Multi-Source Interference Task (MSIT) reaction time for patients with Parkinson’s Disease (PD) with a MOCA ≥ 26 (grey; PD) or < 26 (red; PDCI) under stimulation conditions STN DBS OFF, 4 Hz, and ∼130 Hz. **B**) Interference cost (slower reaction times for incongruent trials) was higher for the PDCI group. * denotes a significant interaction of cognitive status and stimulation condition and # denotes a significant interaction between cognitive status and trial type. Data are from 4896 trials of the MSIT performed by 7 patients with PD without cognitive impairment (grey) and 10 patients with PDCI (red); thin lines represent mean data from a single patient; thick lines represent mean across patients, and shaded area represents standard error of the mean.

Post-hoc comparisons revealed that, when averaged across trial type, cognitively intact participants with PD exhibited faster reaction times in the OFF condition compared to STN DBS at 4 Hz (estimate −0.02, *p* = 0.02), suggesting 4 Hz stimulation slowed down correct responses. In contrast, reaction times were significantly *faster* among participants in the PDCI group during 4 Hz stimulation compared to the OFF condition (estimate *0.02*, *p* = 0.02), and even faster during ∼130 Hz stimulation compared to OFF (estimate 0.07, *p* < 0.0001) and 4 Hz conditions (estimate 0.05, *p* < 0.0001). Together, these data suggest that for PDCI patients, STN DBS progressively speeds reaction times at 4 Hz and ∼130 Hz, similar to the UPDRS-III (Fig 2A)

Next, we used mixed-effects logistic regression to investigate MSIT accuracy. We found main effects of trial type (*χ²*_(1)_ = 23.9, *p* < 0.001) and cognitive status (*χ²*_(1)_ = 5.9, *p* = 0.02). Crucially, cognitive status showed a significant two-way interaction with stimulation condition (*χ²*_(2)_ =16.3, p<0.001), indicating that 4 Hz STN DBS improved accuracy in PDCI but not PD. Because there were no other two-way or three-way interactions including interactions with trial type, we plotted MSIT accuracy as a function of stimulation condition in PD and PDCI patients (Fig 4; see Fig S1 for accuracy as a function of trial type and conflict costs).

**Figure 4:**
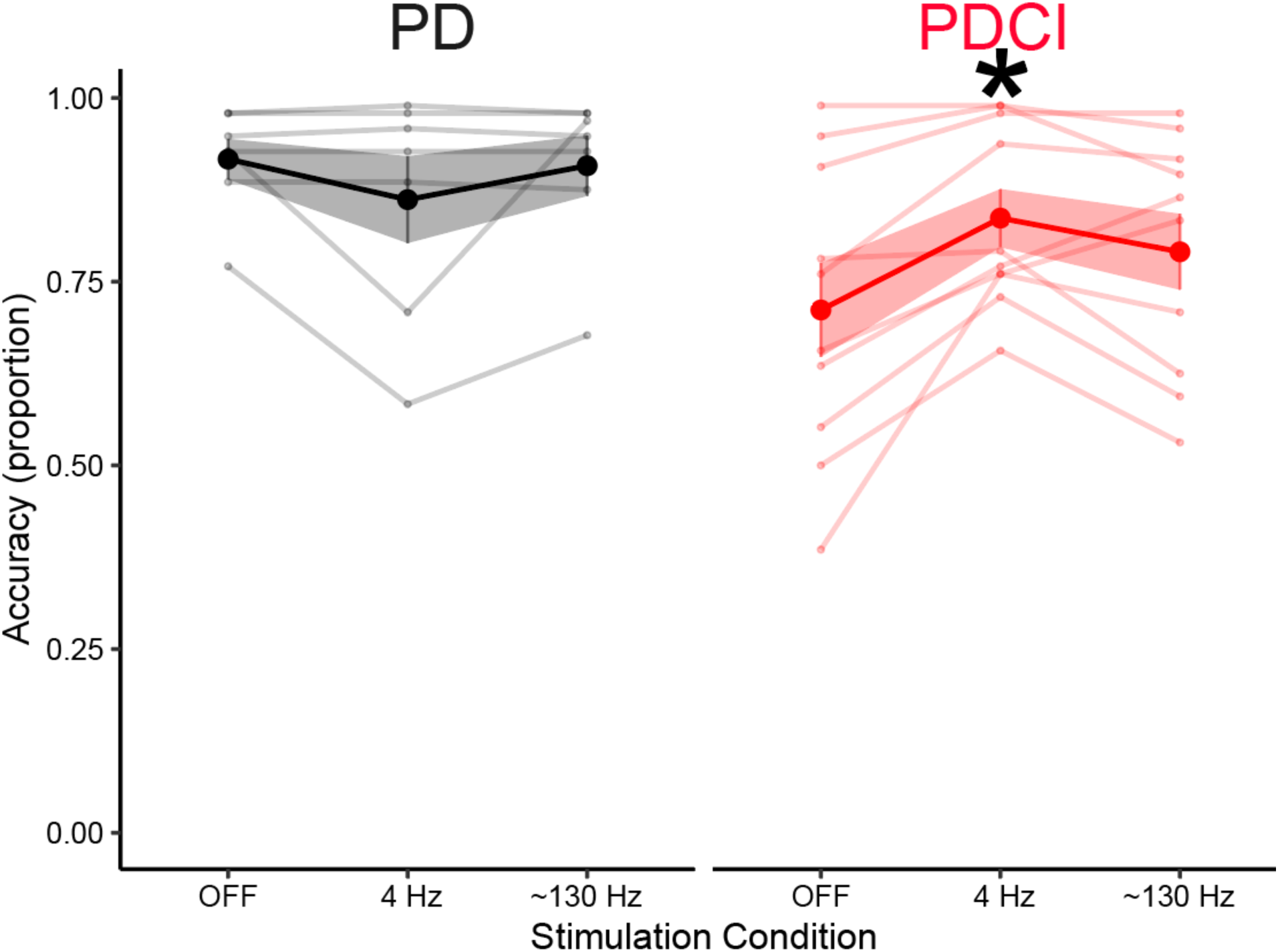
4 Hz Subthalamic Nucleus (STN) Deep Brain Stimulation (DBS) improves MSIT performance accuracy in patients with Parkinson’s Disease (PD) and in PD patients with Cognitive Impairment (PDCI). Multi-Source Interference Task (MSIT) accuracy in patients with PD with a Montreal Cognitive Assessment (MOCA) score ≥ 26 (grey; PD) or < 26 (red; PDCI) under stimulation conditions: STN DBS OFF, at 4 Hz, and ∼130 Hz. * denotes a significant interaction of cognitive status and stimulation condition. Same trials and patients as Fig 2-3; thin lines represent mean data from a single patient; thick lines represent mean across patients, and shaded area represents standard error of the mean.

Post-hoc testing revealed that for patients with PDCI, 4 Hz STN DBS resulted in higher accuracy compared both to DBS OFF (odds ratio = 0.3, *p* < 0.0001) and ∼130 Hz (odds ratio = 1.6, *p* < 0.0001, Fig 2). For patients with PD with normal cognitive function, the effect was opposite: 4 Hz decreased accuracy relative to DBS OFF (odds ratio = 1.8, *p* = 0.01) and relative to ∼130 Hz (odds ratio = 0.5, *p* = 0.01). Together, these data provide robust evidence in support our hypothesis that 4 Hz STN DBS improves MSIT performance by improving accuracy selectively in patients with PDCI (Fig 4). These findings indicate that 4 Hz STN DBS speeds MSIT reaction time and improves MSIT accuracy in PDCI but not PD, and that these effects are distinct from ∼130 Hz stimulation which did not improve UPDRS and reaction-time speed.

## DISCUSSION

In this study, we found 4 Hz STN DBS improves cognitive performance in patients with PDCI. We tested a cognitively diverse group of patients with PD who receive clinical STN DBS by assessing their performance on the MSIT. First, motor function was improved at 4 Hz as measured by the MDS-UPDRS III and was further improved at ∼130 Hz. In line with this, patients with PDCI had progressively decreased MSIT reaction times at 4 and ∼130 Hz. Strikingly, we found that 4 Hz STN DBS, but not ∼130 Hz, improved MSIT performance accuracy. These effects were not observed in PD patients with normal cognition and suggest that in PDCI, 4 Hz STN DBS optimizes responding to maximize MSIT accuracy. Our data provides new evidence that 4 Hz STN DBS improves MSIT performance specifically in patients with PDCI.

PDCI is characterized by attenuated evoked low-frequency frontal midline activity 1-8 Hz in both delta and theta bands, which are linked to cognitive control (Cavanagh and Frank, 2014). These deficits are recapitulated in rodent models that disrupt dopamine (Kim and Narayanan, 2018; Parker et al., 2015; Weber et al., 2025). Indeed, resting-state electroencephalography (EEG) shows that PD patients have increased delta/theta activity (Anjum et al., 2024; Caviness et al., 2016, 2015), but impaired ability to modulate these bands during cognitive tasks (Narayanan et al., 2024; Singh et al., 2023, 2018; Yıldırım et al., 2024). Cortical and STN regions are coherent at ∼4 Hz and are connected by monosynaptic hyperdirect projections (Cole et al., 2022; Haynes and Haber, 2013; Nambu et al., 2002; Walker et al., 2012; Zavala et al., 2016, 2014). This evidence suggests that 4 Hz STN DBS has the potential to modulate cortical networks and enhance cognitive control, which is consistent with the findings from our study.

Our work adds to a growing body of literature that suggests STN DBS at theta frequencies boosts cognitive performance. Previous work by our lab shows that 4 Hz stimulation improves interval timing (Kelley et al., 2018). Similarly, 5 Hz STN DBS improved Stroop task accuracy, and 6 Hz STN DBS improved performance on a working memory task in a frequency-specific manner, whereas stimulation at beta and gamma frequencies did not have the same effect (Salehi et al., 2024; Scangos et al., 2018). Frontal cortical activation is associated with conflict processing deficits, and 5 Hz STN stimulation increased frontal activity and improved conflict resolution (Xie et al., 2025). Moreover, we found that 4 Hz STN DBS caused patients to move slower and be more accurate during Simon-reaction time tasks, supporting prior theoretical models that the STN specifically contributes to raised decision thresholds under conflict (Cavanagh et al., 2011; Cole et al., 2025). Our current findings are also supported by other low-frequency cortical stimulation techniques that boost cognitive control in healthy patients (Demeter, 2016; Gratton et al., 2013; Grover et al., 2023, 2022; Reinhart and Nguyen, 2019). Notably, stimulation at a slightly higher frequency (10 Hz) improved some measures of verbal fluency, although not executive functions (Lam et al., 2021; Ricciardi et al., 2025), further supporting frequency-specific effects of STN DBS.

A major outcome of our current findings is that the beneficial effects of low frequency STN DBS may be maximized in patients with PDCI who presumably have the greatest dysfunction in cortical low-frequency rhythms. This finding is clinically relevant because PDCI patients often are not eligible to undergo STN DBS, yet our work suggests that these patients may experience benefits to both motor and cognitive function with a combination of low and high-frequency stimulation, greatly expanding the utility of DBS in this patient population.

Importantly, 4 Hz STN DBS speeded MSIT reaction time and improved MSIT accuracy but did not have clear effects on cognitive control, as indexed by congruent vs. incongruent MSIT trial types. Our findings are interesting because cognitive control robustly modulates evoked cortical 5-7 Hz theta activity (Cavanagh and Frank, 2014). However, we note that PD patients have more broad-based deficits in cortical rhythms, with increased resting-state delta and theta activity between 1-8 Hz (Anjum et al., 2024; Caviness et al., 2016; Singh et al., 2023) and decreased evoked delta/theta activity that is not specifically related to cognitive control (Singh et al., 2023, 2018). Furthermore, prior low-frequency STN DBS studies have not found specific effects on cognitive control (Salehi et al., 2024; Scangos et al., 2018; Xie et al., 2025). Together, these data support a view where low-frequency activity in PD is modulated by task engagement but not control, and 4 Hz STN DBS boosts these signals, which leads to improved task accuracy. This model could be helpful in designing next-generation neurophysiological markers or neuromodulation therapies for PD.

Although our study revealed important findings, there are several limitations. First, we did not test other frequencies for STN DBS due to the complexity of the MSIT task, time constraints, and fatigue minimization in our participant population. Second, we did not perform a range of neuropsychological tests, thus we were unable to subtype PDCI. For example, MOCA scores in our patient population ranged from 16-27, and included two individuals who scored less than 21 and may be classified as having PDD. Our small sample size did not allow us to investigate PDD, but it is possible that relationship between stimulation and cognitive control differs as cognitive impairment becomes even more severe. Third, it is unclear how MSIT performance translates to functions that are required for the activities of daily life. Finally, we did not perform electrically-stimulated functional MR imaging and we do not have detailed anatomical information for all patients (Vitek et al., 2022; Xie et al., 2025), which could help guide STN stimulation to maximally stimulate prefrontal and cingulate networks and boost cognitive function. Anatomical specificity is an important consideration for future studies, especially as complex programming settings (multiple frequencies across different contacts or leads) could be necessary to maximize benefits for various symptoms. Finally, we note that 4 Hz STN DBS had worse motor function than 130 Hz STN DBS despite improved MSIT performance, indicating that further development, optimization, and testing of low-frequency STN DBS is required for cognitive as well as motor performance.

Collectively, our work shows that 4 Hz STN DBS can improve accuracy during a cognitive control task in patients with PD and impaired cognitive function. Although further rigorous study is required, our findings suggest that low-frequency 4 Hz stimulation boosts cognition in patients with PD and associated cognitive impairment, providing evidence to advance brain stimulation interventions for individuals with neurodegenerative diseases.

## Data Availability

Code and data are available at https://doi.org/10.5281/zenodo.18985728.

## Acknowledgment

We gratefully acknowledge Jennifer Barr of the Scientific Editing and Research Communication Core at the University of Iowa Carver College of Medicine for critical reading of the manuscript, and Patrick Ten Eyck and Linder Wendt from the Biomedical Epidemiology and Research Design Core for statistical review.

## Authors’ Roles

Rachel C. Cole: design, execution, analysis, writing, editing of final version of the manuscript

James F. Cavanagh; design, writing, editing of final version of the manuscript.

Qiang Zhang: execution, editing of final version of the manuscript

Christopher L. Groth: execution, editing of final version of the manuscript

Juan Vivanco-Suarez; execution, analysis, editing of final version of the manuscript

Arturo I. Espinoza: execution, editing of final version of the manuscript

Jeremy D. Greenlee: writing, editing of final version of the manuscript

Nandakumar S. Narayanan: design, analysis, writing, editing of final version of the manuscript

---

**Table S1:** Levodopa and DBS programming characteristics.

| Group | LEDD | MOCA | Right STN |  | Left STN |  | UPDRS |  |  |
| --- | --- | --- | --- | --- | --- | --- | --- | --- | --- |
|  |  |  |  |  |  |  | OFF | 4 Hz | ~130 Hz |
| PD | 550 | 27 | 130Hz | 11+ 10- 0.5V<br>60µs | 130Hz | C+ 3- 0.5V<br>60µs | 9 | 14 | 3 |
| PD | 1243 | 27 | 125 Hz | 9+ 11- 1.95V 60<br>µs | 125<br>Hz | 3+ 2- 1.4V<br>60µs | 25 | 20 | 9 |
| PD | 3150 | 27 | 160 Hz | C- 10- 2.2V<br>90 µs | 160<br>Hz | 3+ 2- 2.4V<br>90µs | 15 | 7 | 11 |
| PD | 2079 | 26 | 140 Hz | 10- 9+ 1.9V<br>90 µs | 140<br>Hz | C+ 2- 0.75V<br>60µs | 41 | 37 | 9 |
| PD | 600 | 26 | 120 Hz | 11+ 10- 4V<br>60 µs | 120<br>Hz | 3 + 1- 3.4V<br>70µs | 59 | 40 | 42 |
| PD | 1380 | 26 | 130 Hz | C+ 9-<br>60 µs | 130<br>Hz | C+ 2- 1.1mA<br>60µs | 23 | 31 | 29 |
| PD | 700 | 26 | 130 Hz | 11+ 9- 3.4V<br>90 µs | 130<br>Hz | 3+ 1- 3.2V<br>70µs | 18 | 22 | 5 |
| PDCI | 900 | 25 | 130 Hz | 11 + 10- 1.6V<br>60 µs | 130<br>Hz | 3+ 2- 2.15V<br>60µs | 36 | 9 | 13 |
| PDCI | 1285 | 25 | 110 Hz | 3 + 2- 4.35V<br>100 µs | 110<br>Hz | 3+ 2- 4.25V<br>150µs | 76 | 60 | 32 |
| PDCI | 650 | 25 | 130 Hz | C+ 10-/9- 3.3V<br>90 µs | 130<br>Hz | C+ 2- 3.2V<br>90µs | 45 | 21 | 19 |
| PDCI | 1980 | 24 | 80 Hz | 11+ 10- 1- 3.45V<br>60 µs | 80 Hz | 3+ 2-/1- 3.45V<br>90µs | 61 | 57 | 45 |
| PDCI | 650 | 24 | 130 Hz | C+ 9- 1.7V<br>60µs | 130<br>Hz | C+ 1- 1.4V<br>60µs | 56 | 38 | 28 |
| PDCI | 935 | 23 | 130 Hz | C+ 11- 2V<br>60 µs | 130<br>Hz | C+ 3- 2V<br>60µs | 62 | 40 | 40 |
| PDCI | 374 | 20 | 130 Hz | C+ 10- 1.5V<br>90µs | 130<br>Hz | 3+ 1-/ 2- 5.6V<br>90µs | 63 | 62 | 31 |
| PDCI | 950 | 18 | 130 Hz | 3+ 1- 1.0V<br>60 µs | 130<br>Hz | 11- 10+ 1.0V<br>60µs | 33 | 23 | 18 |
| PDCI | 1050 | 18 | 150 Hz | C+ 8- 3.8V<br>90 µs | 150<br>Hz | C+ 3- 3.5V<br>80µs | 53 | 49 | 32 |
| PDCI | 0 | 16 | 140 Hz | 11+/10+ 9- 3.9V<br>110 µs | 140<br>Hz | 3+ 2-/1- 3.9V<br>110µs | 60 | 54 | 33 |

LEDD, Levodopa-equivalent dose; MOCA, Montreal Cognitive Assessment; STN, subthalamic
nucleus; UPDRS, Unified Parkinson's Disease Rating Scale, Part III total score; PD, Parkinson's
Disease; PDCI, Parkinson's Disease- Cognitive Impairment

**Figure S1:**
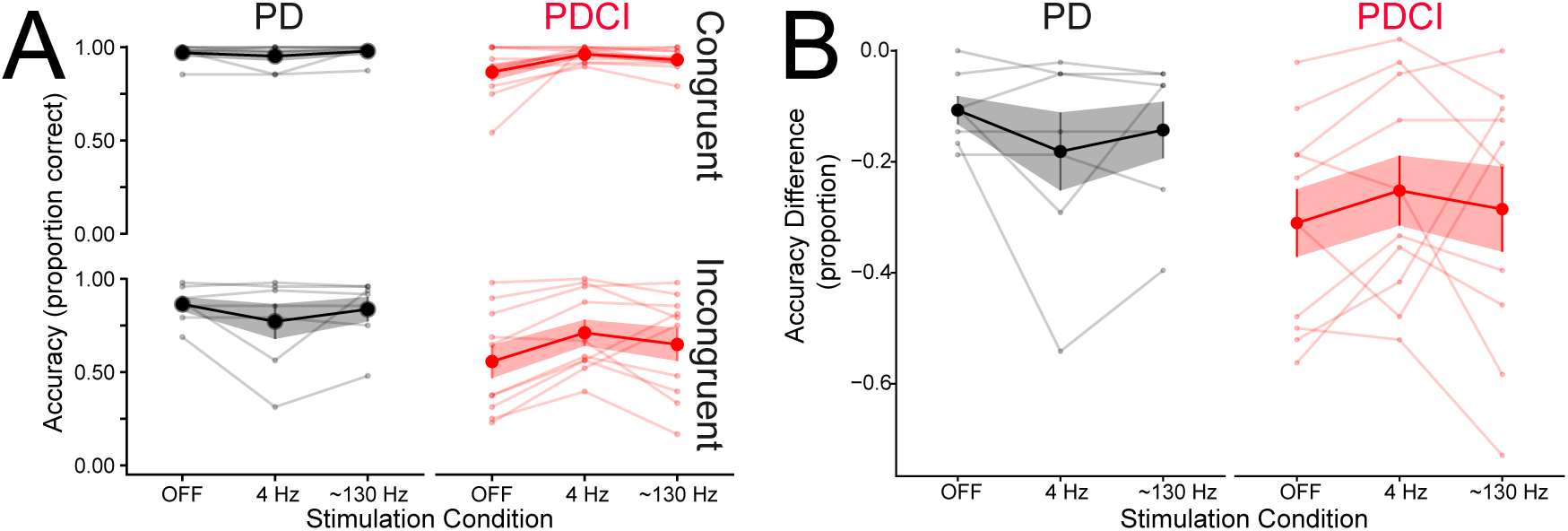
Accuracy in patients with PD and PD Cognitive Impairment (PDCI). **A**) Multi-Source Interference Task (MSIT) accuracy for patients with Parkinson’s Disease with a MOCA ≥ 26 (grey; PD) or < 26 (red; PDCI) under stimulation conditions STN DBS OFF, 4 Hz, and ∼130 Hz for congruent and incongruent trials. **B**) Accuracy interference cost. Data are from 4896 trials of the MSIT performed by 7 patients with PD without cognitive impairment (grey) and 10 patients with PDCI (red); thin lines represent mean data from a single patient; thick lines represent mean across patients, and shaded area represents standard error of the mean.

